# Genetic drivers of etiologic heterogeneity in thyroid cancer

**DOI:** 10.1101/2025.05.15.25327708

**Authors:** Yon Ho Jee, Nikita Pozdeyev, Christopher R. Gignoux, Samantha White, Bryan R. Haugen, Anurag Verma, Raitis Peculis, Vita Rovite, Ashley J Mulford, Alan R Sanders, Cari M Kitahara, Mary Pat Reeve, Peter Kraft, Alicia R. Martin

## Abstract

Thyroid cancer is the most common endocrine malignancy, yet its biological underpinnings remain incompletely understood. We conducted a multi-ancestry genome-wide association study meta-analysis of thyroid cancer (16,167 cases and 2,430,374 controls), identifying 51 independent loci, including 21 novel signals. We analyzed the associations of thyroid cancer risk alleles with 151 other thyroid-cancer-related traits. These pleiotropic relationships reveal mechanistic clusters linked to thyroid function, oncogenic pathways, and mixed physiological function. Two thyroid-specific clusters associated with thyroid stimulating hormone, influencing thyroid growth and function and were enriched in thyroid tissues. Oncogenic clusters included DNA repair (*ATM*, *CHEK2*, *TP53*) and telomere maintenance (*TERT*) genes, implicating shared cancer mechanisms. Cluster-specific polygenic scores were associated with thyroid disease, cancer, and metabolic traits across ancestry groups, suggesting distinct genetic subtypes of thyroid cancer risk. These results demonstrate the utility of pleiotropy-based approaches in uncovering thyroid cancer mechanisms and advancing genetically informed risk stratification.

## Introduction

Thyroid cancer is the most prevalent endocrine malignancy, with an incidence rate of 22.8 per 100,000 per year in women and 8.0 per 100,000 per year in men in the U.S. between 2014 and 2018^1^. The vast majority of thyroid cancers are papillary thyroid carcinomas (PTC) (∼85%), followed by follicular thyroid carcinomas (FTC) (∼10%), while medullary (MTC) (∼2-3%) and anaplastic (ATC) thyroid carcinomas (∼1-2%) are considerably rarer^2^. Prognosis varies substantially by histologic subtype: PTC and FTC, which together comprise the differentiated thyroid cancers arising from thyroid follicular epithelial cells, are associated with favorable outcomes, with high 10-year survival rate (90 to 95%)^3^; in contrast, ATC is highly aggressive, with a median survival of 3-5 months^4^. MTC, which originates from parafollicular C cells, is frequently associated with germline RET mutations in hereditary syndromes such as multiple endocrine neoplasia type 2^5,6^. Thyroid cancer demonstrates moderate heritability, with estimates suggesting genetic factors account for approximately 53% of the disease risk^7^. Globally, thyroid cancer incidence rates have markedly increased in recent decades^8^. While this rise has been largely attributed to improved detection of small, localized slow-growing tumors with high survival rates, the concurrent increase in advanced-stage tumors and thyroid cancer mortality suggests that additional etiological factors contribute to disease progression^9,10^. Established risk factors include childhood exposure to ionizing radiation, while emerging evidence highlights the role of obesity and endocrine-disrupting chemicals in thyroid carcinogenesis^11–13^. However, the underlying biological mechanisms linking these risk factors to thyroid cancer remain poorly understood.

Thyroid function plays a critical role in metabolic homeostasis, and its dysregulation has been linked to benign thyroid disorders, such as goiter and hyperthyroidism, both of which have been associated with an increased risk of thyroid cancer^14^. Additionally, autoimmune thyroid diseases, including Hashimoto’s thyroiditis and Graves’ disease, have been linked to an elevated risk of thyroid cancer, suggesting a potential immune-mediated component in thyroid tumorigenesis^15,16^. Thyroid-stimulating hormone (TSH), a key regulator of thyroid follicular cell growth, has been implicated in thyroid cancer development^17,18^. Genetic variants associated with TSH levels may exert pleiotropic effects on thyroid function through the negative feedback regulation of thyroid hormones, as well as on thyroid cell proliferation, potentially increasing the risk of both goiter and thyroid cancer^19^. However, studies have reported an inverse causal association between TSH levels and thyroid cancer risk, suggesting a more complex interplay between thyroid function and malignancy that warrants further investigation^19–21^. Beyond thyroid hormones, other endocrine factors, including sex hormones, growth hormones, and stress hormones, have been implicated in thyroid tumorigenesis^22^, although the mechanisms underlying the crosstalk between these hormonal systems and their influence on thyroid cancer progression remain unclear^23^.

As genome-wide association studies (GWAS) expand across multiple traits, new opportunities have emerged to leverage cross-trait genetic association patterns to uncover shared disease mechanisms. Studies have demonstrated widespread pleiotropy across the human genome, revealing that genetic variants often influence multiple diseases through shared molecular mechanisms^24^. Pleiotropy dissection provides a powerful framework to cluster genetic associations into biologically meaningful groups, enabling a deeper understanding of how genetic variation drives disease heterogeneity^25^. Recent applications of this approach in type 2 and gestational diabetes^26–29^, autoimmune hypothyroidism^30^, and lung diseases^31,32^ have demonstrated its utility in identifying genetic subgroups that would otherwise be obscured in single-trait GWAS analyses. However, pleiotropy dissection has not yet been applied to thyroid cancer, limiting our ability to differentiate mechanistically distinct genetic pathways underlying its pathophysiology. Given the substantial overlap in genetic susceptibility loci across thyroid diseases^18^, large-scale pleiotropy-informed analyses of thyroid cancer could provide an opportunity to elucidate the complex underlying biological mechanisms of thyroid dysfunction and oncogenesis.

Here, we present findings from a meta-analysis of thyroid cancer GWAS data comprising over 2.5 million individuals of diverse ancestry, more than doubling the effective sample size of prior efforts^17,33–44^. Using cross-trait associations, we characterize the etiological heterogeneity of thyroid cancer and identify genetic clusters that are enriched for distinct tissue- and cell-specific regulatory elements. Furthermore, we construct partitioned polygenic scores (pPGSs) across multiple ancestry groups and assess their associations with thyroid cancer-related outcomes, advancing our understanding of the genetic architecture underlying thyroid cancer.

## Results

### Discovery of thyroid cancer loci

We conducted a fixed-effect GWAS meta-analysis of thyroid cancer using inverse-variance weighted data from 18 biobanks across diverse ancestry groups, comprising 16,167 cases and 2,430,374 controls (Supplementary Table 1). After performing QC, we identified 66 independent thyroid cancer association signals (*P*<5×10^-8^), each represented by an independent index variant (Methods). These 66 association signals mapped to 51 loci, of which 21 (41%) loci have not been previously reported in thyroid cancer GWASs (Supplementary Table 2).

Our top thyroid cancer loci mapped to genes with known functional relevance in thyroid tumorigenesis. The most significant association was observed near *PTCSC2/FOXE1* (rs4273946; beta-C=-0.438, *p*=6.07×10^-235^), a gene previously implicated in thyroid cancer susceptibility^17,45^ (Supplementary Figure 1a). The second most significant variant was located in the intergenic region of *DIRC3* (rs57481445; beta-A=-0.290, *p*=1.34×10^-117^), supporting prior evidence of its regulatory role in thyroid cancer risk^46^. Additional significant intronic variants included rs4733129 in *NRG1* (beta-T=-0.256, *p*=3.68×10^-100^), rs17020141 in *VAV3* (beta-A=-0.188, *p*=6.09×10^-21^), and rs12463278 in *INSR* (beta-A=-0.134, *p*=7.23×10^-15^). These genes are involved in TSH regulation and hypothyroidism, further supporting their potential contribution to thyroid cancer pathogenesis^18,19,34^.

Several thyroid cancer-associated variants overlapped with well-established cancer predisposition genes, reinforcing their role in oncogenic pathways. Notably, rs78378222 at 3’ UTR of *TP53*, rs186430430 (intronic) in *CHEK2*, rs10069690 (intronic) in *TERT*, and rs72743461 (intronic) in *SMAD3* were significantly associated with both thyroid cancer and other malignancies, particularly breast, colorectal, glioma, and prostate cancers^47–51^. Furthermore, a novel missense variant in *ATM* (rs1800057; beta-C=-0.252, *p*=2.89×10^-08^, AF=0.0226) was identified (Supplementary Figure 1b). ATM plays a critical role in the DNA damage response pathway^52^.

Two novel variants may implicate broader endocrine regulatory mechanisms in thyroid cancer susceptibility. For example, the association between rs2842870 (intronic) and thyroid cancer in *PMF1* (beta-T=-0.085, *p*=1.63×10^-12^) was previously also associated with mosaic loss of the Y chromosome (mLOY) (rs2842873 near *PMF1*: beta-T=-0.056, *p*= 1.5×10^-36^ in a cohort of 95,380 Japanese men)^53^. Prior studies have demonstrated that Y-chromosome alterations are linked to increased risk of a range of malignancies that include lung, bladder, and prostate cancers^54^. While thyroid cancer tends to be more aggressive in males^55^, our meta-analysis was not stratified by sex, and further investigation is needed to determine whether sex-specific tumorigenesis mechanisms contribute to risk at this locus. Another intronic variant that is associated with thyroid cancer, rs726686 (non-coding RNA intronic) in *LINC02428* (beta-A=0.082, *p*=1.03×10^-9^, AF=0.1812), was previously associated with sex hormone-binding globulin (SHBG) (beta-A=0.008, *p*=1.1×10^-13^)^56^. SHBG is a key regulator of circulating sex hormones and has been implicated in the etiology of multiple cancers^56,57^; however, its relationship to thyroid cancer risk remains unclear^15,58,59^. Given the pleiotropic nature of SHBG-associated variants^60^ and the lack of consistent directional effects on thyroid cancer risk, this association should be interpreted with caution. Together, these findings suggest broader involvement of endocrine regulatory axes, including thyroidal, gonadal, and adrenal pathways, in thyroid cancer susceptibility^23^.

### Heterogeneity of GWAS Loci Across Thyroid Cancer Subtypes

To characterize the heterogeneity of germline risk across thyroid cancer subtypes, we evaluated 66 lead thyroid cancer variants from the meta-analysis in FinnGen across PTC, FTC, and MTC. In FinnGen, 86% (2534/2963) of thyroid cancers were papillary, 13% (385/2963) follicular, 3% (75/2963) medullary, and 1.7% (49/2963) anaplastic (Supplementary Figure 3). The remaining 5.8% were listed generally as thyroid cancer without a particular designation. PTC and FTC, which together comprise the differentiated thyroid cancers arising from thyroid follicular epithelial cells, demonstrated broadly concordant effects across most loci (Figure 2, Supplementary Table 3). Several loci showed strong and consistent effects in PTC and FTC, including *CHEK2* (rs186430430; PTC beta=□1.29, *p*=1.75×10^-29^; FTC beta=1.02, *p*=0.0020), *EIF4ENIF1* (rs34152346; PTC beta=0.74, *p*=9.20×10^-12^; FTC beta=0.83, *p*=0.03), and *PTCSC2/FOXE1* (rs4273946; PTC beta=-0.46, *p*=8.36×10^-58^; FTC beta=-0.32, *p*=2.04×10^-5^). In contrast, effect sizes in MTC were generally attenuated or absent, consistent with both the distinct cellular origin of MTC from parafollicular C cells and its relatively low case count (n=75), which constrains statistical power. Although *VAV3* (rs17020141) appeared nominally associated with MTC (beta=0.57, *p*=0.029), this association was not observed in the overall thyroid cancer in FinnGen (beta=-0.04, *p*=0.34), and was directionally inconsistent with PTC (beta=-0.077, p=0.092) or FTC (beta=-0.035, *p*=0.77), suggesting that the signal may reflect noise or cohort-specific variation rather than true MTC-specific susceptibility. Conversely, the *TG* locus (rs78775620) showed robust associations in both PTC (rs78775620 beta=□0.33, *p*=1.86×10^-4^) and MTC (beta=1.23, *p*=0.031), but not FTC (beta=-0.01, *p*=0.96), indicating partially overlapping germline influences across histologies. However, given the limited number of MTC cases, these findings should be interpreted with caution and warrant replication in larger, subtype-specific cohorts. In FinnGen, 71% (165/2334) of PTC patients had concurrent diagnoses of FTC, and only 0.7% (16/2334) had overlap with MTC (Supplementary Figure 3).

### Mechanistic clusters of thyroid cancer index variants

In post-GWAS analysis, we next aimed to identify distinct genetic mechanisms driving thyroid cancer using the study design outlined in Figure 1. Genetic correlation analysis with 151 thyroid-cancer-related traits (Methods; Supplementary Table 4) revealed that thyroid cancer shares significant yet moderate genome-wide correlation with several thyroid disorders, immune-related, and cancer phenotypes (significant r_g_ range: -0.24 to 0.53) (Supplementary Table 12, Supplementary Figure 3). Notably, we observed positive genetic correlations with thyroid diseases such as Graves’ disease (r_g_=0.212, *p*=5.43×10^-4^), nontoxic goiter (r_g_=0.427, *p*=0.007), autoimmune hypothyroidism (AIHT) (r_g_=0.197, *p*=0.006), and negative correlations with TSH (r_g_=-0.243, *p*=3.03×10^-4^). Further moderate correlations were observed with type 2 diabetes (T2D) (r_g_=0.074, *p*=0.015), prostate cancer (r_g_=0.120, *p*=0.009), and breast cancer (r_g_=0.127, *p*=0.007).

**Figure 1.**
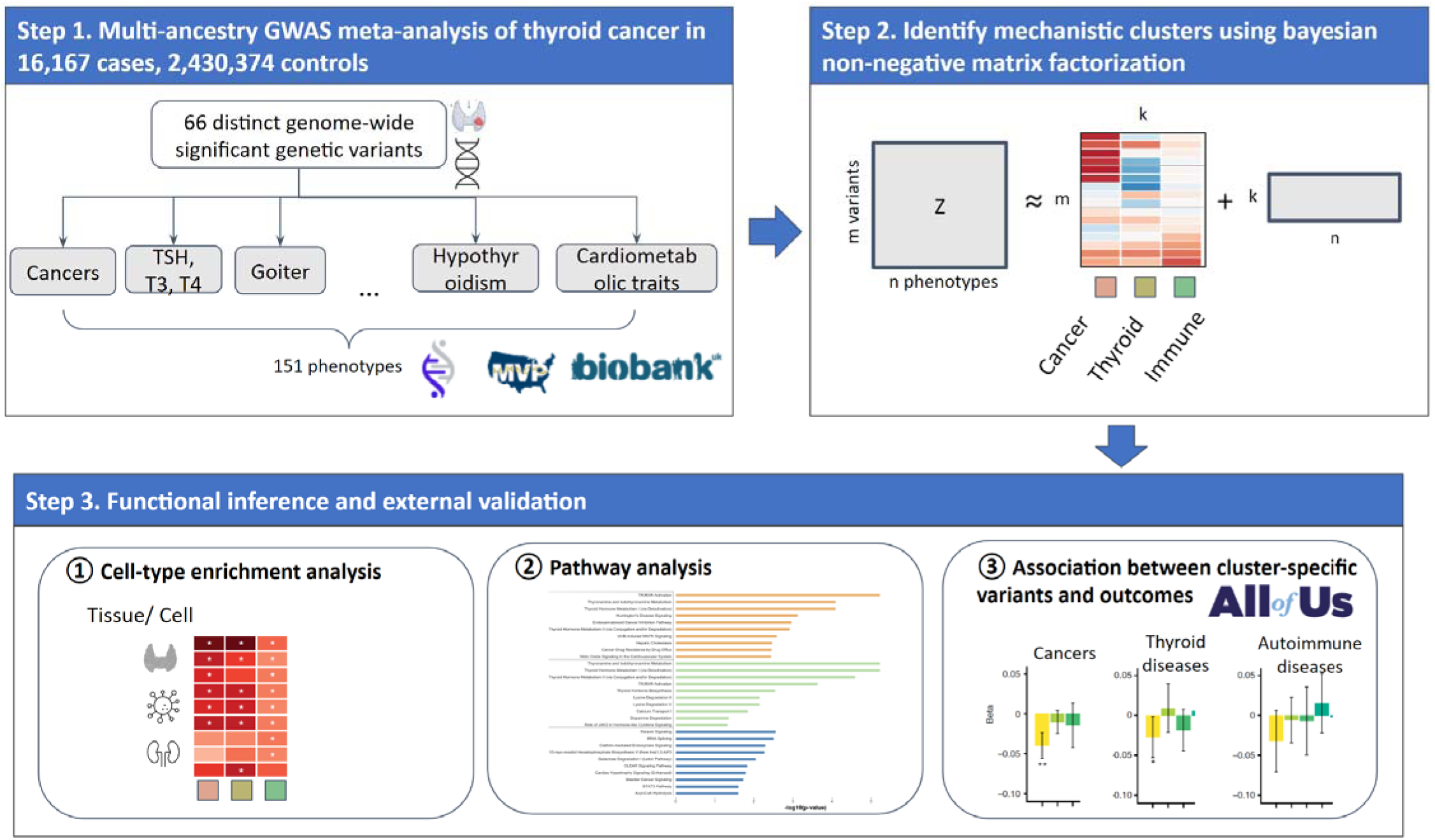
Overview of study design integrating multi-ancestry GWAS, pleiotropy-informed clustering, and functional validation. Schematic of the analytic framework used to identify and interpret germline risk mechanisms in thyroid cancer. Step 1: A multi-ancestry genome-wide association meta-analysis of 16,167 thyroid cancer cases and 2,430,374 controls identified 66 independent genome-wide significant variants. Pleiotropic associations with 151 traits were aggregated across multiple biobanks. Step 2: Bayesian non-negative matrix factorization (bNMF) was applied to the variant–phenotype association matrix to derive mechanistic clusters. Step 3: Functional interpretation of each cluster included (1) cell-type specific enrichment, (2) pathway enrichment analysis, and (3) external replication of cluster-specific genetic risk in the All of Us cohort across disease domains.

Leveraging pleiotropy across these 151 related traits, we used Bayesian non-negative matrix factorization (bNMF) to classify thyroid cancer-associated variants into biologically meaningful clusters. The bNMF analysis identified five mechanistic clusters as providing the best fit to the data (Table 1). As bNMF clusters both genetic variants and phenotypic traits, the top-weighted loci and traits in each cluster may provide insight into their underlying biological mechanisms. The resulting five clusters include one thyroid dysfunction cluster, one thyroid overactivity cluster, two cancer clusters, and one mixed function cluster (Supplementary Table 5).

**Table 1.** Overview of thyroid cancer genetic clusters.

| Cluster (no. of variants) | Key top-weighted traits* | Key top-weighted loci |
| --- | --- | --- |
| 1. Thyroid Gland Dysfunction (12) | TSH(-), Nontoxic multinodular goitre(+), FT3(+), FT4(+), QRS duration(-), Urate(-) | *MAF, CAPZB, VAV3, INSR, MBIP, NRG1, TG, RPSAP52, FOXE1, LINC00609, TGFB2, DIRC3* |
| 2. Thyroid Overactivity (2) | Disorders of the thyroid gland(-), TSH(-), FT4(-), TT3(+), Nontoxic multinodular goitre(-), FT3(+) | *FOXE1, VAV3* |
| 3. DNA Damage Response (9) | Leiomyoma of uterus(+), All cancers(+), SHBG(-), Prostate cancer(+), BMR(+), Skin cancer(+), RBC(+), MRV(-), Height(+), Polycystic ovarian syndrome(+), PP(-), Platelet crit(+), FT4(+), Breast cancer(+) | *TP53, ATM, CHEK2, TTC28, STN1, TERT, EIF4ENIF1, SORCS3* |
| 4. Telomere Maintenance (10) | Leukocyte telomere length(+), Leiomyoma of uterus(+), MRV(-), Platelet crit(+), RBC(+), Eosinophil percentage(-), Neutrophil percentage(+), Total protein(-), Monocyte percentage(-), | *ACTRT3/TERC, STN1, TERT, TYMS, GPR37, ZNF257, SLK, UCKL1, ENOSF1, LOC100287846* |
| 5. Mixed function (3) | Heel BMD(+), QRS duration(+), ALP(+), Platelet distribution width(+), P duration(+), RBC distribution width(+), TG(-), Neuroticism score(-), eGFR(+), AST(+), Eosinophil percentage(+), ALT(+), GGT(+), MAP(+), Total protein(+) | *PRAG1, CEP120, SMAD3* |

\*+/-: thyroid cancer risk alleles associated with increased or decreased phenotype values.
ALP, Alkaline phosphatase; ALT, Alanine aminotransferase; AST, Aspartate aminotransferase; BMR, basal metabolic
rate; BP, blood pressure; BMD, bone mineral density; eGFR, Estimated glomerular filtration rate; FT3, free T3; GGT,
Gamma glutamyltransferase; MAP, Mean arterial pressure; Mean reticulocyte volume, MRV; PP, Pulse pressure; RBC,
Red blood cell; SHBG, sex hormone binding globulin; TG, Triglycerides; TSH, thyroid stimulating hormone; TT3, total T3.

Clusters 1 (Thyroid Gland Dysfunction) and 2 (Thyroid Overactivity) reflect mechanisms related to thyroid function. Both clusters include decreased TSH and increased free triiodothyronine (FT3), indicating a hyperthyroid state. Thyroid cancer risk alleles linked to Cluster 1 are positively associated with nontoxic multinodular goiter, whereas risk alleles linked to Cluster 2 show a negative association. Both clusters contain variants near *VAV3* and *FOXE1*, genes with established roles in thyroid carcinogenesis via the MAPK and PI3K pathways^61,62^. Additionally, Cluster 1 includes variants near *NRG1*, *INSR*, and *TGFB2*, suggesting interactions between growth factor signaling and epithelial-mesenchymal transition (EMT) in thyroid tumor progression^63,64^. Collectively, this cluster likely captures structural thyroid changes associated with hormone overproduction, which may contribute to thyroid autonomy and tumor aggressiveness. Interestingly, Cluster 1 is also characterized by shortened QRS duration, which may reflect electrocardiogram features associated with overt hyperthyroidism^65–67^.

Thyroid cancer risk alleles linked to Clusters 3 (DNA Damage Response) and 4 (Telomere Maintenance) are strongly associated with cancers and tumorigenic pathways. Both clusters show a strong correlation with leiomyoma of the uterus, but they differ in other specific cancer associations. Cluster 3 is linked to all-cancer risk, prostate cancer, skin cancer, and breast cancer, while Cluster 4 is associated with higher IGF-1 levels, lower eosinophil percentage, and higher neutrophil count. Cluster 3 (DNA Damage Response) contains variants mapped to key tumor suppressors such as *TP53*, *CHEK2*, and *ATM*, all of which play central roles in DNA damage repair pathways^47,68^. *CHEK2* germline mutations have been implicated in breast, colorectal, and thyroid cancers, and loss of function mutations in *CHEK2* with compromised checkpoint-mediated DNA repair, increasing genomic instability^48^. Similarly, both germline and somatic mutations in *ATM* impair the cell cycle checkpoint response, leading to an increased risk of malignancy^69^. Cluster 4 (Telomere Maintenance) features germline variants near *TERT* and TERC/*ACTRT3*, genes previously linked to telomere length regulation in malignancies^70^. Somatic mutations in the TERT promoter are major determinants of increased telomerase activity, with variants linked to multiple cancers, including thyroid cancer^71,72^. Additionally, the cluster includes *STN1*, a component of the CST (CTC1-STN1-TEN1) complex, which is critical for telomere replication and stability^73^, further supporting the cluster’s role in telomere maintenance and oncogenesis.

Thyroid cancer risk alleles linked to Cluster 5 (Mixed Function) are strongly associated with various sets of traits, including heel bone mineral density, QRS and P duration, alkaline phosphatase, triglycerides, and mean arterial pressure. Although this cluster includes variants near *SMAD3*, which is a key transcription factor in TGF-β signaling, with broad roles in development, fibrosis and cancer progression^74^, the lack of common biology with the other top-weighted genes, such as *PRAG1* and *CEP120*, limits biological interpretation, and the cluster remains mechanistically uncharacterized.

To assess the robustness of these mechanistic clusters, we conducted a sensitivity analysis restricted to 73 expert-curated traits (Methods; Supplementary Table 4). The resulting four clusters recapitulated the core biological mechanisms observed in the main analysis, including two thyroid-related clusters, one cancer-associated cluster, and one mixed function cluster, with strong overlap in top-weighted traits and loci (Supplementary Table 10). For example, Cluster 1, enriched for TSH, FT3, and nontoxic multinodular goiter, corresponds to the original Thyroid Gland Dysfunction cluster. Similarly, the cancer-focused cluster preserved defining features of the DNA Damage Response and Telomere Maintenance cluster, retaining key loci such as *TP53*, *CHEK2*, and *TERT*. Cluster 4 in the sensitivity analysis mirrors the original Mixed Function cluster, showing enrichment for traits including QRS and P duration, pulse pressure, and mean arterial pressure. The consistent emergence of these clusters across distinct input data support the stability of the underlying biological structure and suggest the clustering results are not overly sensitive to trait selection.

### Regulatory processes underlying clusters

To further assess the mechanistic associations with each cluster, we examined the epigenomic enrichment of top loci using cell-type-specific annotations (Figure 2, Supplementary Table 6). We assessed whether variants within each cluster show significant overlap with genes expressed in specific tissues or with chromatin accessibility markers specific to a particular tissue^75^ (Methods).

**Figure 2.**
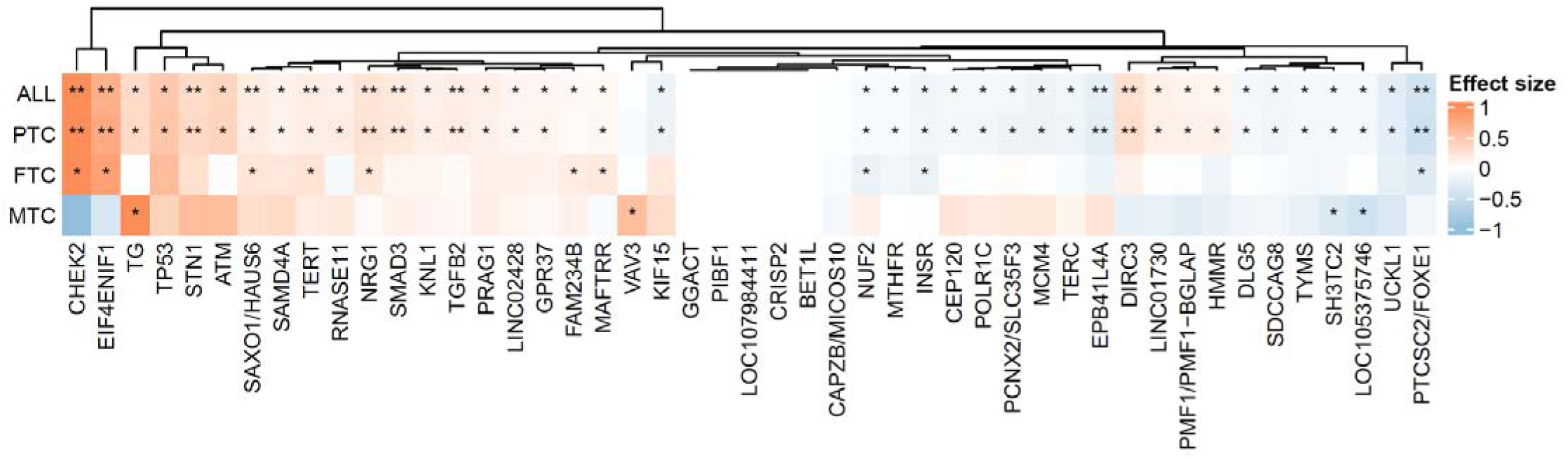
Subtype-specific effect sizes for thyroid cancer risk variants. Heatmap of beta coefficients for 51 loci across thyroid cancer subtypes in FinnGen: papillary (PTC), follicular (FTC), and medullary (MTC). Each column represents a locus, and rows correspond to subtype-specific effect sizes. Variants are clustered by similarity in effect profiles. p□<□0.05 (*), p□<□5×10□□ (**).

The Thyroid Gland Dysregulation (*P*=0.019 for Thyroid_gland_H3K27ac) and Thyroid Overactivity clusters (*P*<0.001 for Thyroid_gland_H3K27ac and H3K4me1) showed significant enrichment in thyroid gland cells, as identified in EN-TEx chromatin accessibility data^76^. However, gene expression data from Franke et al^77,78^ and GTEx data did not show significant enrichment (Figure 3, Supplementary Table 6).

**Figure 3.**
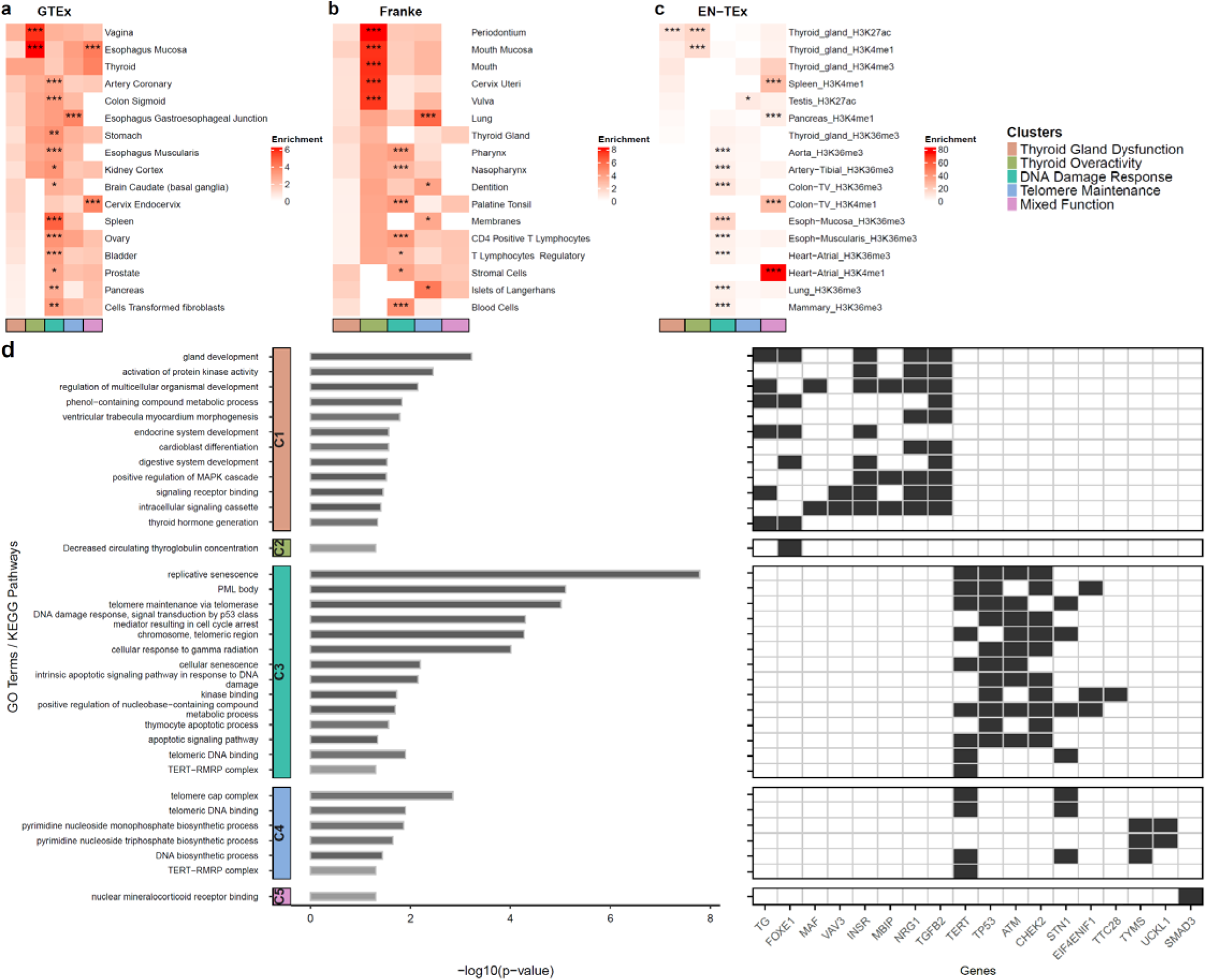
Functional enrichment of cluster-specific thyroid cancer risk loci across tissues, cell types, and biological pathways. **a-c.** Heatmaps display enrichment patterns across top-weighted loci from each cluster across tissues or cell types. Only tissues or cell types with significant enrichment are displayed. For each dataset, the top 17 tissues or cell types are shown, selected based on their average enrichment across clusters with at least one cluster-level enrichment (PlJ<lJ0.05). Thyroid tissue is included for reference, even if not significantly enriched in all clusters. (a) Selected tissues from 53 total in GTEx. (b) Selected tissues from 152 total in the Franke dataset. (c) Selected cell-type and histone mark combinations from four activating histone marks analyzed in EN-TEx. \*\*\**P* < 0.001, \**P* < 0.05. **d.** Pathway enrichment of the top-weighted loci from each cluster. Left bars represent multiple-testing adjusted p-values of gene set analysis for pathways identified within each cluster. Colored squares represent locus involvement in the corresponding pathway.

For the other clusters, enrichment was not restricted to a single tissue type, which is expected given that these pathways regulate cell cycle, metabolic processes, and systemic hormone signaling across multiple tissues. This widespread enrichment pattern suggests that genetic variants in these clusters likely contribute to disease susceptibility through diverse regulatory effects beyond thyroid-specific mechanisms.

Pathway enrichment analysis of genes near index variants in each cluster identified relevant biological pathways (Figure 3, Supplementary Table 7). In the Thyroid Gland Dysfunction cluster, key loci, including *FOXE1*, *INSR*, *TGFB2*, *TG*, and *NRG1*, were significantly enriched in pathways related to gland development (*P*=6.00×10^-4^) and regulation of multicellular organismal development (*P*=0.007). These findings recapitulate their established roles in thyroid proliferation and differentiation^79^. The Thyroid Overactivity cluster demonstrated enrichment for the pathway associated with decreased circulating thyroglobulin concentration, primarily driven by *FOXE1* (P□=□0.05), further supporting its functional relevance in thyroid hormone biology. As expected, the DNA Damage Response cluster showed enrichment in pathways associated with DNA damage response, signal transduction by p53 class mediator resulting in cell cycle arrest (*P*=5.00×10^-5^), cellular response to gamma radiation (*P*=9.76×10^-5^), and intrinsic apoptotic signaling pathway (*P*=0.007), primarily driven by *TP53*, *CHEK2*, and *ATM*^47,68^. Similarly, the Telomere Maintenance cluster was enriched for telomere cap complex (*P*=0.001), telomeric DNA binding (*P*=0.013), and TERT-RMRP complex (*P*=0.05), reinforcing its role in maintaining telomere integrity in tumorigenesis^71^.

For the Mixed function cluster, nuclear mineralocorticoid receptor (MR) binding was the only significantly enriched pathway (*P*=0.05), driven exclusively by *SMAD3*. Although MR signaling has been implicated in cancer biology, including cellular proliferation, EMT, and metabolic regulation across various cancer types^80^, the enrichment is based on a single gene, and the top-weighted genes in the cluster do not converge on a shared biological pathway. As such, we refrain from assigning a pathway-based label to this cluster. Instead, the cluster appears to be characterized by its leading phenotypic associations, heel bone mineral density, QRS duration, and alkaline phosphatase, without evidence for a shared underlying biological mechanism.

### Associations of partitioned polygenic scores (pPGSs) with outcomes

To validate our findings, we examined whether partitioned polygenic scores (pPGS) derived from the top-weighted loci in each cluster were associated with a range of traits in an independent sample. We focused on thyroid cancer-related phenotypes available in the All of Us (AoU) Research Program^81^ (Figure 4, Supplementary Table 8-9).

**Figure 4.**
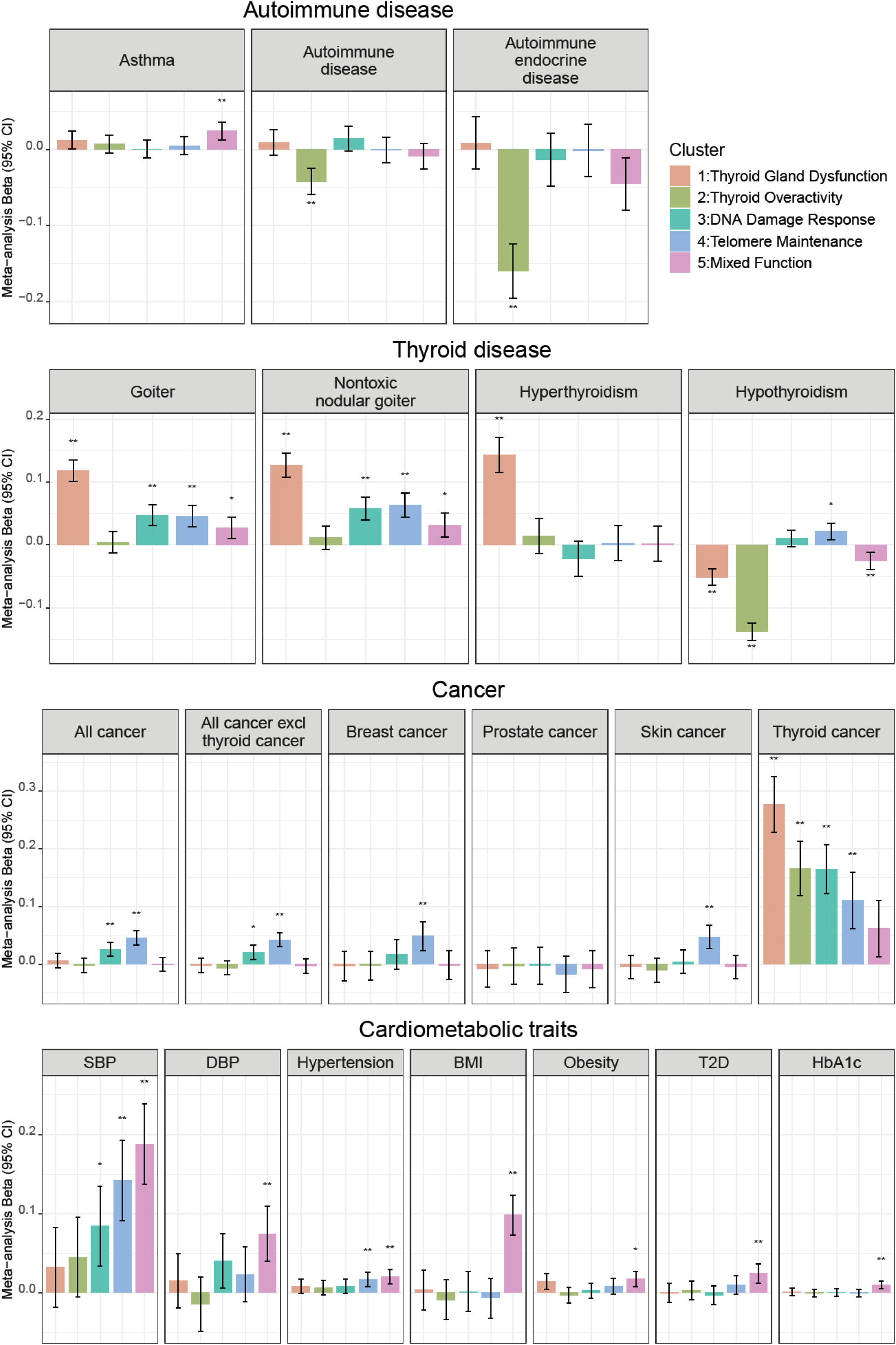
Associations of cluster-specific partitioned polygenic score (pPGS) with thyroid cancer-related outcomes in AllofUs data. Summaries of the associations of each cluster-specific component of the pPGS with autoimmune disease, thyroid disease, cancer, and cardiometabolic traits. The height of each bar corresponds to the log-odds ratio (beta) per standard deviation of the pPGS, and the grey bar shows the 95% confidence interval. *P < 0.05/20, Bonferroni correction for 20 outcomes; **P < 0.05/(5*20), Bonferroni correction for 20 outcomes and 5 clusters. Note: y-axis ranges differ across disease categories to reflect varying effect size distributions.

The pPGS from the Thyroid Gland Dysfunction cluster showed strongest associations with increased risk of goiter (Beta=0.119, *P*=5.91×10^-43^), nontoxic goiter (Beta=0.127, *P*=4.31×10^-39^), and hyperthyroidism (Beta=0.144, *P*=9.13×10^-24^). Conversely, this cluster was associated with a decreased risk of hypothyroidism (Beta=-0.051, *P*=1.10×10^-13^), consistent with the hyperthyroid phenotype linked to genetic variants within this cluster.

Similarly, the Thyroid Overactivity cluster was significantly associated with a lower risk for hypothyroidism (Beta=-0.138, *P*=1.79×10^-85^), autoimmune disease (Beta=-0.042, *P*=1.02×10^-6^), and autoimmune endocrine disease (Beta=-0.159, *P*=1.47×10^-18^). In the AoU data browser, Hashimoto thyroiditis comprises the majority of autoimmune endocrine disorder diagnoses.

As expected, our strongest associations that served as a positive control were between all five thyroid cancer clusters and thyroid cancer risk in AoU, with effect estimates ranging from Beta=0.062, *P*=1.42×10^-2^ for the Mixed Function cluster to Beta=0.277, *P*=3.30×10^-29^ for the Thyroid Gland Dysfunction cluster.

pPGS of cluster-specific variants also exhibit specificity in trait associations; only the DNA Damage Response and Telomere Maintenance clusters were significantly associated with other cancers. These clusters were positively associated with all cancers excluding thyroid cancer (Beta=0.020, *P*=0.001 for the DNA Damage Response; Beta=0.043, *P*=1.87×10^-11^ for the Telomere Maintenance cluster). Specific associations were observed for the Telomere Maintenance cluster, which was linked to breast cancer (Beta=0.049, *P*=0.0002) and skin cancer (Beta=0.047, *P*=5.76×10^-6^). These findings suggest that these cancer-related clusters extend beyond thyroid cancer and may influence multiple malignancies.

We next evaluated whether pPGS from thyroid cancer clusters were associated with cardiometabolic traits. We found positive associations between systolic blood pressure and multiple clusters, including the Telomere Maintenance (Beta=0.142, *P*=4.59×10^-8^) and Mixed function clusters (Beta=0.188, *P*=4.06×10^-13^). However, only the Mixed Function cluster showed significant associations with various cardiometabolic traits, including diastolic blood pressure (Beta=0.074, *P*=2.26×10^-5^), BMI (Beta=0.098, *P*=2.36×10^-14^), obesity (Beta=0.017, *P*=6.27×10^-4^), type 2 diabetes (T2D) (beta=0.024, *P*=5.74×10^-5^), and HbA1c (beta=0.010, *P*=4.85×10^-5^). These results suggest that the Mixed Function cluster may reflect shared genetic components between thyroid cancer and metabolic traits, potentially related to obesity-linked pathways, although the precise biological mechanisms remain to be elucidated.

## Discussion

In this study, we conducted a meta-analysis of thyroid cancer GWASs across diverse ancestry groups, identifying 51 genome-wide significant loci, 21 (41%) of which are novel. By increasing sample size and incorporating more diverse populations, we identified 57% more loci than the largest prior thyroid cancer GWAS from GBMI^33^. Using pleiotropy dissection, we classified 66 independent thyroid cancer-associated variants across 151 traits into five distinct genetic clusters, each representing biologically meaningful pathways. Our findings suggest that thyroid cancer susceptibility is driven by multiple independent mechanisms, including thyroid gland dysfunction, thyroid overactivity, DNA repair deficiency, telomere maintenance, and mixed physiological function.

Previous studies have demonstrated the utility of pleiotropy dissection in complex diseases by defining genetic subgroups associated with distinct biological mechanisms. Suzuki et al. aggregated T2D GWAS data from over 2.5 million individuals, including >400,000 cases across multiple ancestry groups (∼40% non-European), identifying 1,289 association signals across 611 loci, including 145 novel loci^28^. Their use of k-means clustering on T2D and 37 cardiometabolic traits revealed eight genetic clusters, enriched in open chromatin regions of pancreatic islets and other cell types, highlighting key regulatory pathways underlying T2D subtypes. Complementary work by Smith et al. applied soft clustering to T2D meta-analysis of 228,000 cases (1.4 million total individuals), integrating 110 T2D-related traits, and identified 12 genetic clusters that largely overlapped with Suzuki et al clusters and were enriched in specific single-cell regulatory regions^27^. By dissecting pleiotropy among thyroid cancer and related traits for the first time, our study demonstrates that distinct genetic clustering not only captures heterogeneity in thyroid function but also links to broader oncogenic, and metabolic pathways, thereby advancing our understanding of thyroid cancer subtypes and precision medicine approaches.

The Thyroid Overactivity and Thyroid Gland Dysregulation clusters highlight how distinct axes of thyroid function may contribute to thyroid cancer susceptibility through different physiological mechanisms. Both clusters capture TSH-driven thyroid function-related pathways but diverge in their phenotypic associations, particularly with goiter. These clusters recapitulate the potential pleiotropic effects of TSH-associated variants, where Thyroid Overactivity cluster primarily reflects feedback regulation of thyroid hormones on the pituitary, while Thyroid Gland Dysregulation cluster is linked to thyroid growth and an increased risk of thyroid cancer and goiter^19^. Reeve et al. previously characterized the genetic architecture of AIHT, identifying both thyroid-specific and immune-mediated genetic contributors using linemodels^82^, an alternative method for dissecting pleiotropic effects across pairs of traits into variants that influence both traits from trait-specific effects^30^. Notably, *FOXE1* and *VAV3*, present in both of our thyroid clusters, showed unique effects in AIHT that are absent in broader autoimmune disease in their analysis, consistent with our findings of their thyroid-specificity. In our subtype-specific analysis, *FOXE1* demonstrated consistent associations with differentiated thyroid cancers (PTC and FTC), but not with MTC, whereas *VAV3* was significantly associated only with MTC, likely reflecting limited statistical power. This subtype-specificity may reflect differences in cell-type-specific gene expression and signaling networks, with *FOXE1* crucial for follicular epithelial cell differentiation and function^83^, while *VAV3* is predominantly involved in RET-mediated signaling pathways characteristic of MTC^61,84^. However, further studies are needed to validate the role of these genes across cancer subtypes. Moreover, *FOXE1* and *TG*, two key loci of the Thyroid Gland Dysregulation cluster, overlap with congenital hypothyroidism risk genes identified in the Genomics England clinical panel^85^, reinforcing their roles in thyroid development and thyroid hormone production. Moreover, several genes involved in both AIHT and TSH regulation (*DIRC3*, *NRG1*, *INSR*, and *VAV3*) in Reeve et al align with the Thyroid Gland Dysfunction cluster. These findings suggest that genetic variation in thyroid-related pathways may influence both autoimmune thyroid diseases and thyroid cancer susceptibility, providing insights into shared molecular mechanisms underlying thyroid dysfunction and tumorigenesis. The Thyroid Gland Dysfunction cluster is also characterized by associations with shortened QRS duration^65–67^. Although even mildly altered thyroid status can affect heart rhythm^86^ and rate^87^, the frequent occurrence of cardiac manifestations in hyperthyroid patients may result from thyrotoxicosis itself, pre-existing heart disease exacerbated by hyperthyroidism, or specific hyperthyroidism-induced cardiac abnormalities^88^. While QRS duration is less sensitive to heart rate than the QT interval^89^, these associations may suggest a potential influence of thyroid hormone dysregulation on cardiac conduction. However, as GWAS of these ECG traits used in this study are not adjusted for heart rate, the mechanisms underlying these associations remain unclear and warrant further investigation.

Beyond TSH-related mechanisms, our discovery of cancer-related clusters (DNA Damage Response and Telomere Maintenance) suggests that thyroid cancer shares fundamental tumorigenic mechanisms with other malignancies. These clusters reflect biologically distinct processes, including genomic instability and replicative immortality, both hallmarks of cancer. The DNA Damage Response cluster includes top loci associated with DNA repair pathways, such as *ATM*, *CHEK2*, and *TP53*. ATM and ATR kinases phosphorylate p53 following DNA damage, regulating cell cycle arrest, DNA repair, and apoptosis; thus, mutations in this cluster may increase genomic instability and predispose individuals to aggressive thyroid cancer subtypes^90^. Similarly, the enrichment of *TERT*-associated loci in the Telomere Maintenance cluster underscores the role of telomere length regulation in thyroid tumorigenesis. Genetic predisposition to telomere maintenance is thought to facilitate continuous cell proliferation and the accumulation of oncogenic mutations, a well-established feature of aggressive thyroid cancers^71^. Notably, these mechanistic overlaps reflect epidemiological patterns of increased co-occurrence between thyroid cancer and other malignancies, particularly breast and kidney cancers, which can be explained by shared genetic and environmental risk factors and medical surveillance^91,92^. While these clusters may not immediately alter clinical management of thyroid cancer, they offer valuable insights into the underlying biological heterogeneity of the disease. Specifically, they may help identify subsets of patients at higher risk of aggressive disease, inform drug repurposing efforts targeting DNA repair or telomerase pathways, and guide future studies on common mechanisms across multiple cancer types. More broadly, these findings illustrate how pleiotropic clustering can reveal latent axes of dysregulation that contribute to disease pathogenesis and may eventually support precision oncology approaches.

The Mixed Function cluster suggests possible metabolic contributions to thyroid cancer risk. This cluster is associated with several cardiometabolic and inflammatory markers and showed enrichment for nuclear MR binding pathways. MR is expressed in both normal thyroid tissues and PTC cells, where its activation by aldosterone induces inflammatory responses such as interleukin-6 expression and modulates thyroid-specific genes^93^. Furthermore, MR expression progressively declines in more aggressive thyroid cancer histotypes, suggesting a potential role in tumor progression^80,93^. Although MR activation is not directly linked to cardiometabolic disease, its pro-inflammatory activity and transcriptional regulation in thyroid cells highlight a potential mechanistic link between hormonal signaling, inflammation, and metabolic dysregulation in thyroid cancer. Further studies are needed to elucidate the precise role of MR signaling within the broader network of metabolic and inflammatory pathways in thyroid cancer. As obesity is a well-established risk factor for several cancers, our findings further support the association between metabolic dysregulation and thyroid tumorigenesis^11^. Importantly, this cluster may also serve as a potential target for primary prevention strategies, such as weight loss or metabolic interventions, in genetically susceptible individuals at increased thyroid cancer risk via cardiometabolic pathways, particularly those related to obesity and T2D. Early lifestyle or pharmacologic interventions could help mitigate disease risk and guide personalized preventive strategies, especially in populations with high prevalence of metabolic syndrome.

Our external validation analysis demonstrated that pPGSs from thyroid, cancer, and cardiometabolic genetic clusters were significantly associated with risks of thyroid disease, cancers, and cardiometabolic conditions, consistent with the key traits identified within each cluster. These findings suggest that genetic clustering may facilitate the stratification of individuals with a genetic predisposition to thyroid cancer into clinically meaningful subgroups, potentially guiding personalized treatment approaches. For instance, somatic mutations in the *TERT* promoter are associated with poor prognosis and loss of radioiodine (RAI) avidity when co-occurring with *BRAF V600E* mutations^94,95^. However, it remains unclear whether germline TERT variations confer similar clinical implications. In cases of RAI-refractory differentiated thyroid cancer, multi-kinase inhibitors such as lenvatinib and sorafenib have been approved for treatment. These agents are typically employed in advanced RAI-refractory disease when other options are limited^96–98^. Similarly, *TP53* mutations are frequently observed in ATC, the most aggressive thyroid cancer subtype^99^. While *TP53* mutations are often linked to treatment-resistant cancers, they have also been proposed as potential biomarkers of immunotherapy responsiveness, as p53 loss has been linked to increased tumor mutational burden, which may enhance response to immune checkpoint inhibitors^100–102^. However, the predictive value of *TP53* mutations for immunotherapy response in thyroid cancer remains uncertain^103^. Future studies should investigate whether patients stratified by genetic clusters exhibit differential responses to targeted therapies to further refine personalized treatment strategies in thyroid cancer.

There are several limitations to this study. First, we were unable to assess the impact of genetic clusters across thyroid cancer histologic types or by stage at diagnosis, which are associated with prognosis and treatment response. Consequently, some observed associations may reflect vertical pleiotropy and reverse causation. For instance, thyroidectomy, a standard treatment for thyroid cancer, induces hypothyroidism, necessitating levothyroxine therapy to suppress TSH levels, potentially leading to iatrogenic hyperthyroidism. This clinical intervention could influence phenotypic associations captured by our thyroid-related clusters. Second, heterogeneity in screening practices and diagnostic criteria across countries, cohorts, and calendar years^104,105^ may contribute to variability in the direction and magnitude of observed associations. Our findings highlight subtype-specific genetic associations, underscoring the need for subtype-stratified studies to further elucidate these genetic relationships. Third, we did not account for prior thyroid disease diagnoses, hormone replacement therapy, or thyroid cancer treatments, all factors that could impact genetic associations^22^. While our results suggest that certain genetic clusters may inform differential outcomes or treatment strategies, their direct clinical application requires further validation. In parallel to our present study, the GBMI has explored the genetic architecture of thyroid diseases and developed clinically relevant polygenic risk score discriminating thyroid cancer and benign thyroid nodules, which may aid in improving diagnosis accuracy of thyroid cancer (Pozdeyev *et al.* 2025 *in prep*). Fourth, our analysis incorporated both expert-curated traits directly relevant to thyroid cancer and additional traits associated indirectly through genetic loci. To assess the potential bias arising from selective trait inclusion, we performed a sensitivity analysis restricted to 73 expert-curated traits, confirming that the robustness of the identified thyroid, cancer, and mixed physiological clusters (Supplementary Table 10). Fifth, we used bNMF for variant clustering; however, alternative methods such as independent component analysis^106^ or linemodels^82^ could yield different partitions of genetic signals. Further validation is needed to assess cluster robustness across analytical approaches.

Our findings demonstrate the value of cross-trait genetic associations in elucidating distinct molecular pathways underlying thyroid cancer. Future research should focus on integrating functional genomics, transcriptomics, and clinical interventions to further refine genetically informed strategies for thyroid cancer management and personalized treatment.

## Methods

### Meta-analysis of thyroid cancer

Following analysis plans outlined previously^33^, each study conducted a GWAS of thyroid cancer using logistic mixed model with SAIGE^107^ or whole genome regression with REGENIE^108^ (Supplementary Table 1). Phenotype definitions for thyroid cancer varied across contributing biobanks, including diagnoses based on ICD-10 codes, phecodes, or self-reported data (Supplementary Table 1). Thus, the analyses included both prevalent and incident thyroid cancer cases, and were not restricted to first cancer occurrences. We conducted a fixed-effect meta-analysis of thyroid cancer using inverse-variance weighting,^109^ combining GWAS summary statistics from two meta-analyses (the GBMI meta-analysis^33^ and a separate meta-analysis of FinnGen R12^110^ and MVP^37^) and four individual biobanks (the the Korean Cancer Prevention Study-II Biobank^111,112^, Latvian National Biobank^113^, Penn Medicine BioBank, and Genomic Health Initiative) (Supplementary Table 1).

To assess heterogeneity across the six sets of GWAS summary statistics contributing to our meta-analysis, we calculated Cochran’s Q statistics. We retained variants present in at least one of the GWAS in this meta-analysis for locus discovery. Given the limitations of high-resolution fine-mapping methods in the context of meta-analyses particularly when performed using different genotyping and imputation techniques, we adopted a conservative strategy for defining lead variants. Specifically, we flagged only the most significantly associated variant within each confidently defined, linkage disequilibrium (LD)-independent locus. LD independent associations were determined as follows: starting with the most significant association with p <5×10^-8^ on each chromosome, we scanned a ±2 Mb window around each genome-wide significant signal. A dynamic LD threshold was then applied, defined as T = min (0.1, r_5_) where r_5_ corresponds to the r^2^ value at which the expected residual chi-square statistic equals 5.0. Secondary associations within the window were retained only if they were genome-wide significant and had r^2^ < T with any more significant signal. In instances where the lead association was exceptionally strong (resulting in T < 0.02), we excluded secondary signals within ±1 Mb to mitigate the risk of false independence due to instability in low r² estimates. We used Finnish LD reference panel sisu4.2.

A locus was classified as previously reported if it contained a lead variant within a ±500 kb flanking window of a known association for the same Experimental Factor Ontology (EFO) term in the Open Target Genetics database (release 22.09)^114^. Otherwise, it was categorized as novel (Supplementary Table 2). To comprehensively identify prior associations, we performed an exhaustive search in the GWAS Catalog and then excluded loci from the list of novel hits if they were reported in the GWAS catalog but not in OpenTargets. Additionally, as FinnGen R12^110^ and MVP^37^ results were not available in the Open Target Genetics database or the GWAS Catalog at the time of evaluation for the novel association, we additionally excluded loci where a ±500 kb flanking window contained any previously reported associations from FinnGen or MVP.

### Defining clusters of thyroid cancer index variants using bNMF

To classify thyroid cancer-associated variants into biologically meaningful subgroups, we applied Bayesian non-negative matrix factorization (bNMF)^26,115^. The clustering was based on GWAS summary results for the independent thyroid-cancer index variants across 151 phenotypes, including 73 expert curated traits associated with risk of thyroid cancer (Thyroid disease, Autoimmune disease, Cancer, Cardiometabolic, Hormone measurement), alongside 78 additional traits linked to categories associated with thyroid cancer variants (Anthropometry, Lung, Mental disease, Biomarker, Hematological traits, Kidney, Reproductive system, Genomic measurement). We prioritized multi-ancestry GWAS for ancillary trait summary statistics; however, when unavailable for a given trait, European-based GWASs were used. Linkage disequilibrium score regression (LDSC) ^116^ was applied to estimate cross-trait genetic correlations (r_g_) between our thyroid cancer meta-analysis and 151 clustering phenotypes.

We applied a modified bNMF pipeline^26,27^. Detailed descriptions of the pipeline are provided in Supplementary Data. Traits were excluded if their median sample size was below 5,000 or if their minimum P value for the final variant set did not meet Bonferroni significance (Pmin > 0.05/66 variants). Additionally, we removed highly correlated traits based on Pearson correlation coefficient (> 0.80) calculated from GWAS summary statistics across index variants, prioritizing traits with the strongest variant–trait associations, resulting in a final set of 100 traits (Supplementary Table 5).

We generated standardized effect sizes for variant trait associations from GWAS by dividing the estimated regression coefficient beta by the standard error, which had been aligned to the thyroid cancer risk-increasing alleles. To account for differences in sample size across studies and enable a more uniform comparison of phenotypes across studies, we scaled the standardized effect sizes by the square root of the study size, as estimated by mean sample size across all SNPs, forming the variant-trait association matrix **Z** (66 variants × 100 traits). Missing variant–trait associations were replaced with zero. For sensitivity analysis, we imputed missing associations using z-scores from proxies (LD r^2^ > 0.5) where possible and otherwise used the median observed standardized effect size for the trait (Supplementary Table 11). The bNMF algorithm was implemented in RStudio and executed for 10,000 iterations with varying initial conditions. The most frequently observed K across 50 chains of 10,000 iterations was selected as the final model. To assess the robustness of the clustering results, we conducted a sensitivity analysis restricting the phenotypic input to the 73 expert-curated traits (reduced to 50 traits after filtering) (Supplementary Table 10).

### Enrichment of thyroid cancer associations for cell-type-specific regions of open chromatin within clusters

To assess whether cluster-specific variants exhibit significant overlap with cell-type-specific regulatory elements, we computed excess overlap as defined in a previous study^117^. Excess overlap quantifies whether two binary annotations co-occur more often than expected by chance. Specifically, for two binary annotation vectors (annot1 and annot2) across *M* variants, it is calculated as:

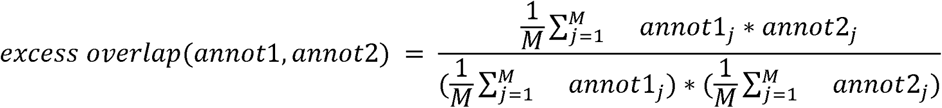

For each cell-type annotation, we define annot1 as a binary vector across the *M* variants, where each entry takes the value 1 if the corresponding variant is a thyroid cancer index variant for a given cluster, and 0 otherwise. Index variants were assigned to a cluster if their cluster weight was above 1.057, a threshold determined to maximize the signal-to-noise ratio, as previously described^26^. Similarly, annot2 is a binary vector indicating whether each of the *M* variants is annotated as enriched in the given cell-type (1) or not (0). This calculation is mathematically equivalent (up to a simple transformation) to computing the Pearson correlation between the two binary annotations.

We applied this metric to five binary cell-type annotation datasets used in LD Score regression with Specifically Expressed Genes (LDSC-SEG)^75^. Gene expression-based annotations (GTEx^118^, Franke^77,78^, ImmGen^119^) were generated by identifying genes with highly tissue- or cell-type-specific expressions (top 10% by specificity metrics) and defining SNP annotations as windows (±100 kb) around these genes. Chromatin-based annotations from Roadmap^120,121^ and EN-TEx datasets^76^ were defined using experimentally identified peaks for DNase I hypersensitivity and activating histone marks (H3K27ac, H3K4me3, H3K4me1, H3K9ac and H3K36me3). For each cluster of thyroid cancer index variants, we mapped them to the full set of *M* SNPs covered by each LDSC-SEG annotation dataset and computed the excess overlap with each cell-type-specific annotation.

The statistical significance of annotations in each cluster was assessed using a permutation test. For each cluster, we permuted annot1 values (thyroid cancer index variant indicators) and recalculated the excess overlap using shuffled labels. After 10,000 permutations, we compared the observed excess overlap to the permuted background using a one-tailed test to determine the significance of each annotation. We corrected for multiple tests and considered statistically significant enrichment at q-value thresholds of 0.1 and 0.001, using Bonferroni correction (for 53 tissues in GTEx, 152 tissues in Franke, 292 immune cell types in ImmGen, 396 in Roadmap, 93 in EN-TEx).

### Pathway Enrichment Analysis

To identify biological pathways enriched in each genetic cluster, we used gProfiler^122^ (g:GOSt) (https://biit.cs.ut.ee/gprofiler/gost) to test for KEGG, WikiPathways, or REACTOME pathway enrichment. g:GOSt conducts functional profiling of gene lists by analyzing various types of biological evidence, performing statistical enrichment analysis to identify over-representation of information from Gene Ontology (GO) terms, biological pathways, regulatory DNA elements, human disease gene annotations, and protein-protein interaction networks.

The functional enrichment of the input gene list is evaluated using a cumulative hypergeometric test, evaluating multiple functional terms simultaneously for each gene list. To control for false positives, we applied g:SCS (Set Counts and Sizes) multiple testing correction, the default method in g:GOSt^123^. g:SCS accounts for overlapping functional terms, making it more conservative than Benjamini-Hochberg False Discovery Rate but less stringent than Bonferroni correction.

### Cluster-specific partitioned polygenic score analyses in AllofUs

To validate the thyroid cancer genetic clusters, we constructed partitioned polygenic scores (pPS) and tested their associations with potential thyroid cancer-related traits in the All of Us Research Program (AoU) database^81^. Phenotypic data were derived from two primary domains in AoU: the conditions domain, which captures medical diagnoses and health outcomes based on standardized clinical codes (e.g., ICD, SNOMED), and the measurement domain, which includes quantitative clinical values resulting from examinations or tests.

The pPGS were calculated as weighted sums of genetic variants within each cluster, with weights derived from the bNMF clustering results. Only variants with cluster weights above 0.9554 were included in the pPGS^26^. The pPGS were standardized to have a mean of zero and unit variance and analyzed separately for four ancestry groups (AMR, AFR, EUR, and EAS) in AoU.

Within each ancestry, association testing was performed using logistic regression for binary traits and linear regression for continuous traits, adjusting for age, sex, and the first ten principal components (PCs). A fixed-effects inverse-variance-weighted meta-analysis was conducted to combine ancestry-specific estimates, with Bonferroni correction applied for the number of outcomes and for both the number of outcomes (20) and the number of clusters examined (5×20).

## Data Availability

The meta-analysis GWAS summary statistics will be made publicly available prior to publication. Data sources for other publicly available GWAS summary statistics are available in Supplementary Table 4.

## Acknowledgements

We want to acknowledge the participants and investigators of all participating biobanks. We are grateful to Heiko Runz for helpful discussions.

This work was supported by a stipend from the PhD Program in Population Health Sciences at Harvard University and the Intramural Research Program of the National Cancer Institute, US National Institutes of Health (NIH). The computations in this paper were run on the FASRC Cannon cluster supported by the FAS Division of Science Research Computing Group at Harvard University.

## Supplementary Figures

**Supplementary Figure 1.**
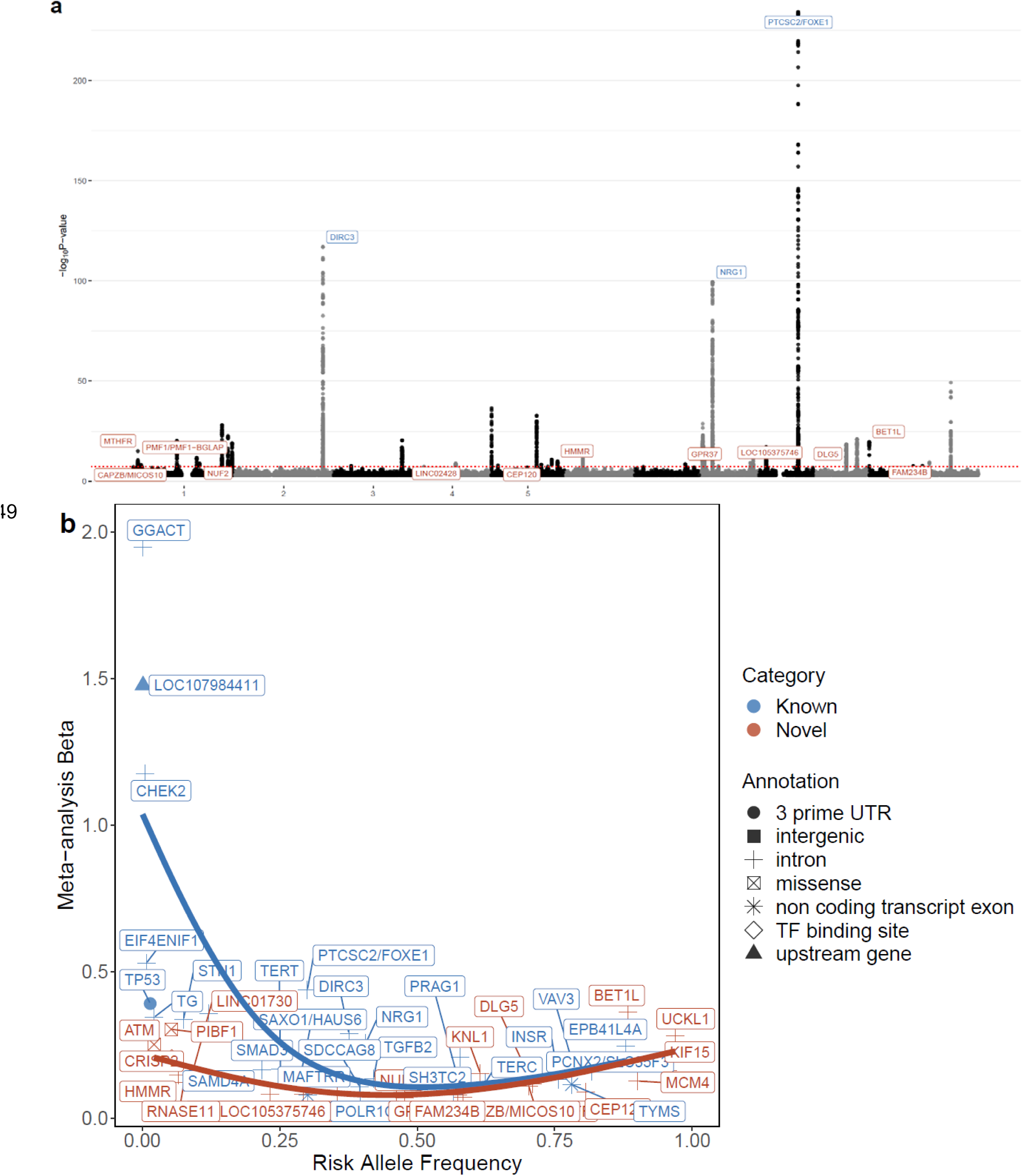
Manhattan plot for the meta-analysis GWAS of thyroid cancer (a) and comparisons of the frequency of thyroid cancer risk allele versus the effect sizes for all 51 thyroid cancer genetic loci (b). Novel thyroid cancer loci are highlighted in red.

**Supplementary Figure 2.**
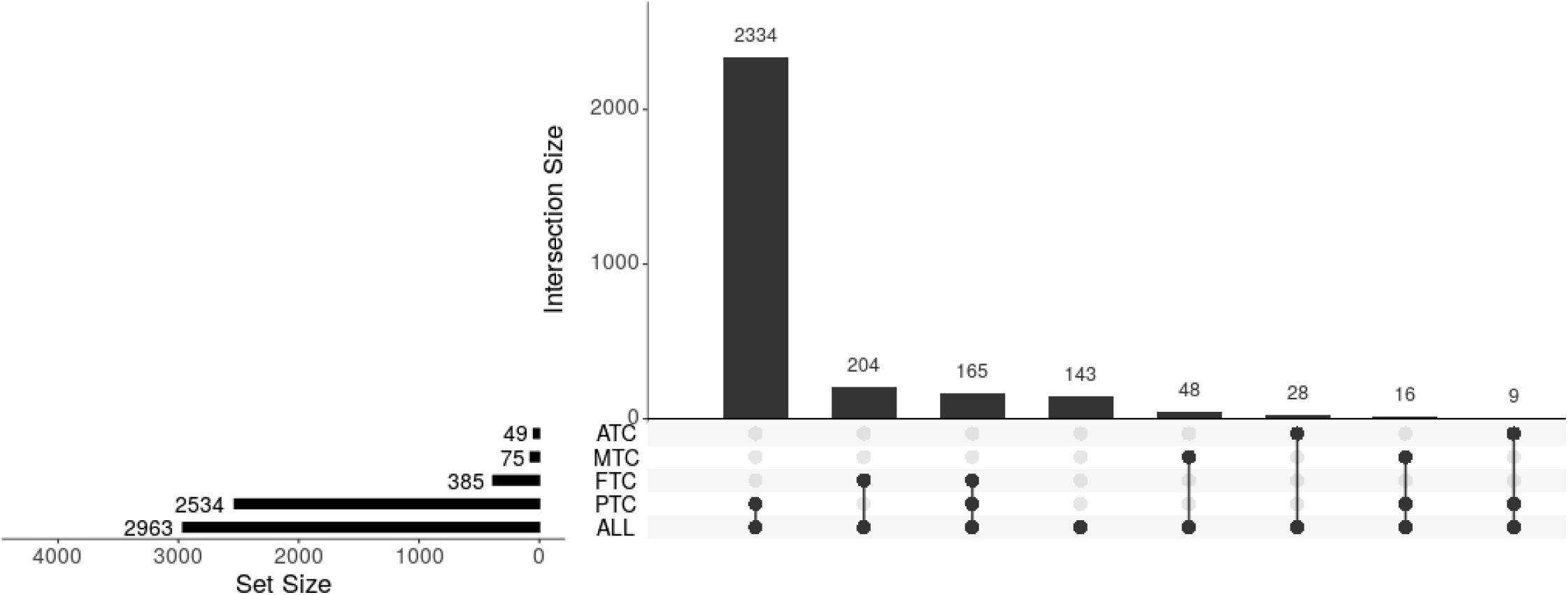
Number of thyroid cancer cases across subtypes in FinnGen. UpSet plot illustrating the intersection of thyroid cancer subtypes defined in FinnGen. The majority of individuals were diagnosed exclusively with papillary thyroid carcinoma (PTC; n = 2,334), followed by follicular thyroid carcinoma (FTC; n = 385), medullary thyroid carcinoma (MTC; n = 48), and anaplastic thyroid carcinoma (ATC; n = 28). A smaller subset of individuals had multiple subtype diagnoses, including concurrent PTC and FTC (n = 165), PTC and MTC (n = 16), and PTC and ATC (n = 9).

**Supplementary Figure 3.**
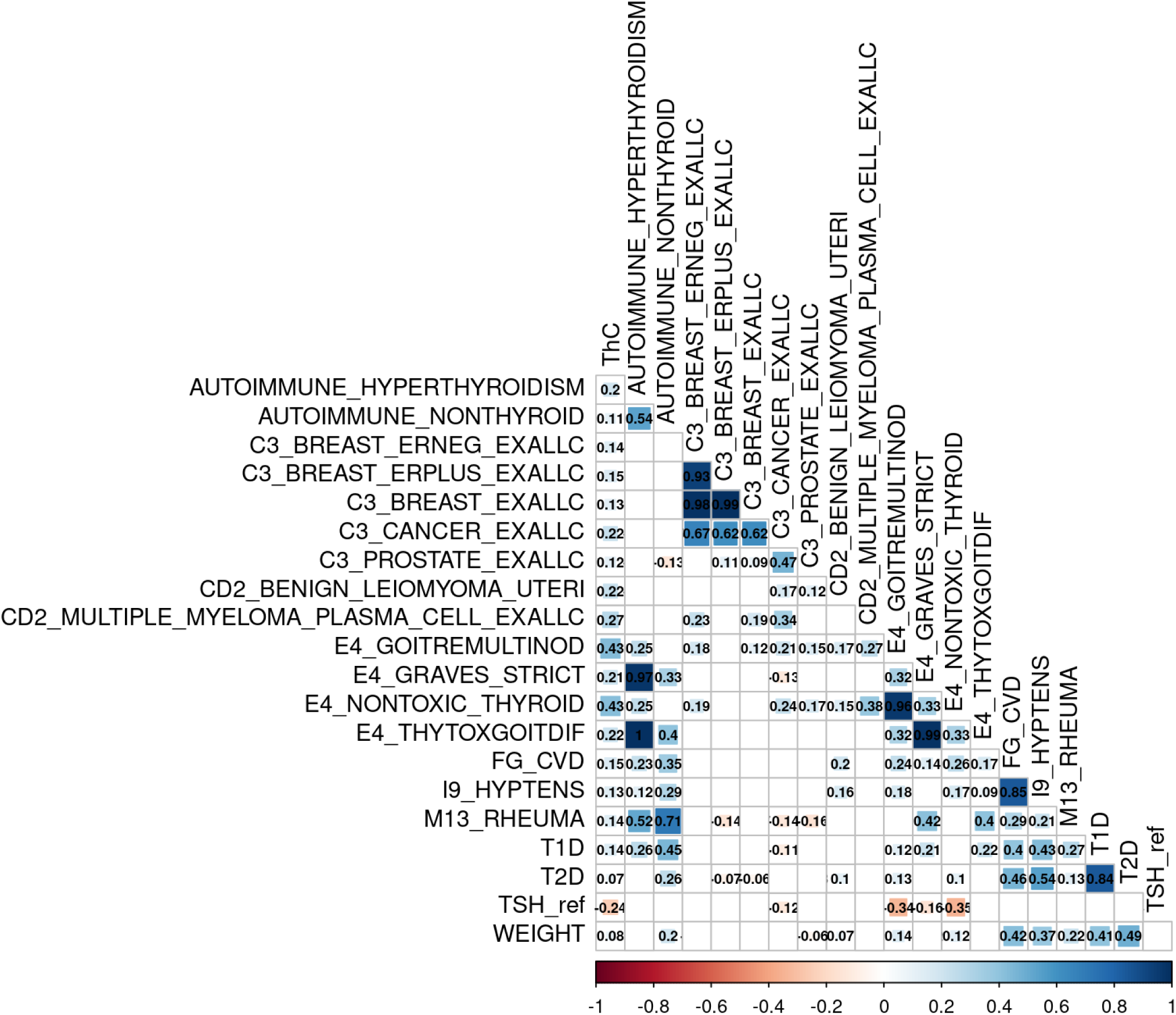
Genetic correlation estimates between thyroid cancer and 151 clustering phenotypes. Only significant genetic correlation estimates with thyroid cancer are presented. Full genetic correlation results are shown in Supplementary Table 12. ThC=thyroid cancer.

